# A Pilot Interventional Study on Feasibility and Effectiveness of the CUE1 device in Parkinson’s disease

**DOI:** 10.1101/2024.10.11.24315305

**Authors:** Viktoria Azoidou, Kira Rowsell, Ellen Camboe, Kamalesh C. Dey, Alexandra Zirra, Corrine Quah, Thomas Boyle, David Gallagher, Alastair J Noyce, Cristina Simonet

**Author notes:** Corresponding authors: Centre for Preventive Neurology, Wolfson Institute of Population Health, London, United Kingdom. &.

## Abstract

**Introduction:** Current treatments for patients with Parkinson’s disease (PwP) can fail to address gait disturbance and falls, which in turn affect quality of life (QoL). The CUE1 device delivers cueing with vibrotactile stimulation showing potential to alleviate motor symptoms and reduce falls based on preliminary user testing results.

**Objectives:** To evaluate the feasibility, safety, and tolerability of CUE1 and its effect on motor and non-motor symptoms in PwP.

**Methods:** PwP used the CUE1 for 9-weeks and were assessed at week 0, 3, 6, and 9 on MDS-UPDRS Part-III, Timed Up and Go (TUG), TUG with dual task (DT), Functional Gait Assessment (FGA), and Bradykinesia Akinesia Incoordination (BRAIN) test. Patients reported outcomes through MDS-UPDRS Part-I, -II, and -IV and Pittsburgh Sleep Quality Index (PSQI).

**Results:** Ten PwP (5 females, age range: 46-80; disease duration: 3-9 years) completed the CUE1 intervention with 100% compliance and no adverse events. CUE1 comfort and usability were rated highly (80%). Immediate CUE1 effect was observed on MDS-UPDRS-III(45.40±12.22 vs 39.60±11.74, p=0.008), TUG(11.53±1.92 vs 11.08±1.94, p=0.022), TUG DT(18.57±5.75 vs 17.61±6.28, p=0.037) and FGA(16.40±3.86 vs 18.60±3.92, p=0.007). Cumulative effect was noted on MDS-UPDRS-III(45.40±12.22 vs 27.80±12.32, p=0.005), FGA(18.60±3.92 vs 23.10± 2.85, p<0.001), TUG DT(18.57±5.75 vs 13.58±7.05, p=0.031), BRAIN kinesia (45.10±14.39 vs 42.10±12.74, p<0.001) and incoordination (24331.09±38017.46 vs 14059.91± 9030.96, p=0.016) scores, PSQI(10.10±4.95 vs 6.90±3.81, p=0.002), MDS-UPDRS-I(18.60±6.75 vs 12.20±3.68, p=0.011), MDS-UPDRS-II(17.30±7.29 vs 11.90±8.67, p=0.002), and MDS-UPDRS-IV(7.50±3.75 vs 3.40±2.95, p=0.003).

**Conclusion:** In this unblinded, feasibility study, cueing with the CUE1 appeared to be a feasible and safe intervention for PwP improving motor and non-motor features.

**PLAIN ABSTRACT:** *Background and Objectives:* This study looked at the use of a device called CUE1, which provides rhythmic pulsing vibration to help reduce movement and non-movement symptoms in people with Parkinson’s disease (PwP). Even with current treatments, issues with walking and falling still affect the quality of life for many patients. The goal of this study was to see if CUE1 could be a helpful, safe, and easy-to-use tool for improving movement symptoms, like walking and risk of falling, as well as non-movement symptoms, like sleep.

*Procedures and Results:* Ten people with Parkinson’s (5 women, aged 46-80) who had Parkinson’s for 3-9 years used the CUE1 device for 9 weeks. Everyone finished the study, using the device regularly with no negative side effects. The device was rated 80% for comfort and ease of use. Significant improvements were seen in movement abilities (using the MDS-UPDRS-III test), walking speed, and fall risk. Over the 9 weeks, participants showed further progress in movement, balance, and coordination. Non-movement symptoms, like sleep, also improved.

*Conclusions:* The CUE1 device, which provides gentle vibrations to assist with movement, was found to be safe, easy to use, and effective. It improved both movement-related symptoms like balance and walking, as well as non-movement symptoms like sleep. This makes it a promising new treatment option for people with Parkinson’s.

## INTRODUCTION

Parkinson’s disease (PD) is a progressive neurodegenerative disorder affecting millions of people worldwide^1^. Motor symptoms include tremor, bradykinesia, rigidity, postural instability, freezing of gait (FOG), and falls, which impair psychological wellbeing, independence, and health-related quality of life (QoL)^2^. Non-motor symptoms, such as sleep issues, sensory impairment, anxiety, and depression, can also be debilitating^2^. The mainstay of treatment for PD is pharmacological therapies as first line, followed by more advanced therapies such as deep brain stimulation as symptoms progress. Both pharmacological and surgical therapies bring their own challenges and can exacerbate postural instability, FOG, and falls, increasing morbidity and mortality and diminishing the QoL in patients with Parkinson’s (PwP)^3–6^. Exercise can improve balance, but managing gait disturbance and preventing falls is challenging due to limited access to specialised physiotherapists^7^ and difficulties with medication adherence^4,8^.

Cueing, a non-invasive technique using external prompts like visual, auditory, or tactile signals, helps manage motor symptoms without invasive intervention^9,10^. However, many cueing methods are not practical for home use, emphasising the need for wearable solutions^9^. Focused vibrotactile stimulation—using localised skin vibrations—has shown consistent benefits for balance and gait issues, and reducing FOG, in contrast to mixed results from whole-body stimulation^11,12^. Vibrotactile stimulation using devices like CUE1^13^, tactile anklets^14^, and Equistasi^12^, shows promise for improving motor symptoms, balance, and gait, and reducing falls. These methods have practical advantages over visual and auditory cues in everyday use^9,10^.

User testing results have demonstrated that the CUE1 improved gait speed as measured by the Timed Up and Go (TUG) test^13^, motor performance on Movement Disorder Society-Sponsored Revision of the Unified Parkinson’s Disease Rating Scale (MDS-UPDRS) Part-III, gait disturbance including FOG^15^, and reduced falls by 83%^16^. With this pilot study we aimed to evaluate the feasibility of the CUE1 in PwP and assess its safety, tolerability, and impact on both motor and non-motor symptoms, including gait, balance, and fall risk. A positive outcome would warrant further exploration of its potential to enhance movement and QoL in PwP.

## METHODS

### Trial Design

This 9-week interventional feasibility study was conducted by staff at Queen Mary University of London (QMUL), UK (reference number: 23/PR/1526; clinical.trials.gov ID: NCT06174948). The study adhered to the Declaration of Helsinki. All patients provided written informed consent before data collection.

### Participants

From March to August 2024, patients were recruited from the Neurology Department at Barts Health NHS and the Care of the Elderly Department at Homerton University Hospital NHS Trust. Assessment took place at the Centre for Preventive Neurology, Wolfson Institute of Population Health, QMUL. Two neurologists identified potential participants (CS and AJN), and a senior neurological physiotherapist (VA), trained for this study, managed consent and data collection. See Figure 1 for the study flowchart.

**Figure 1.**
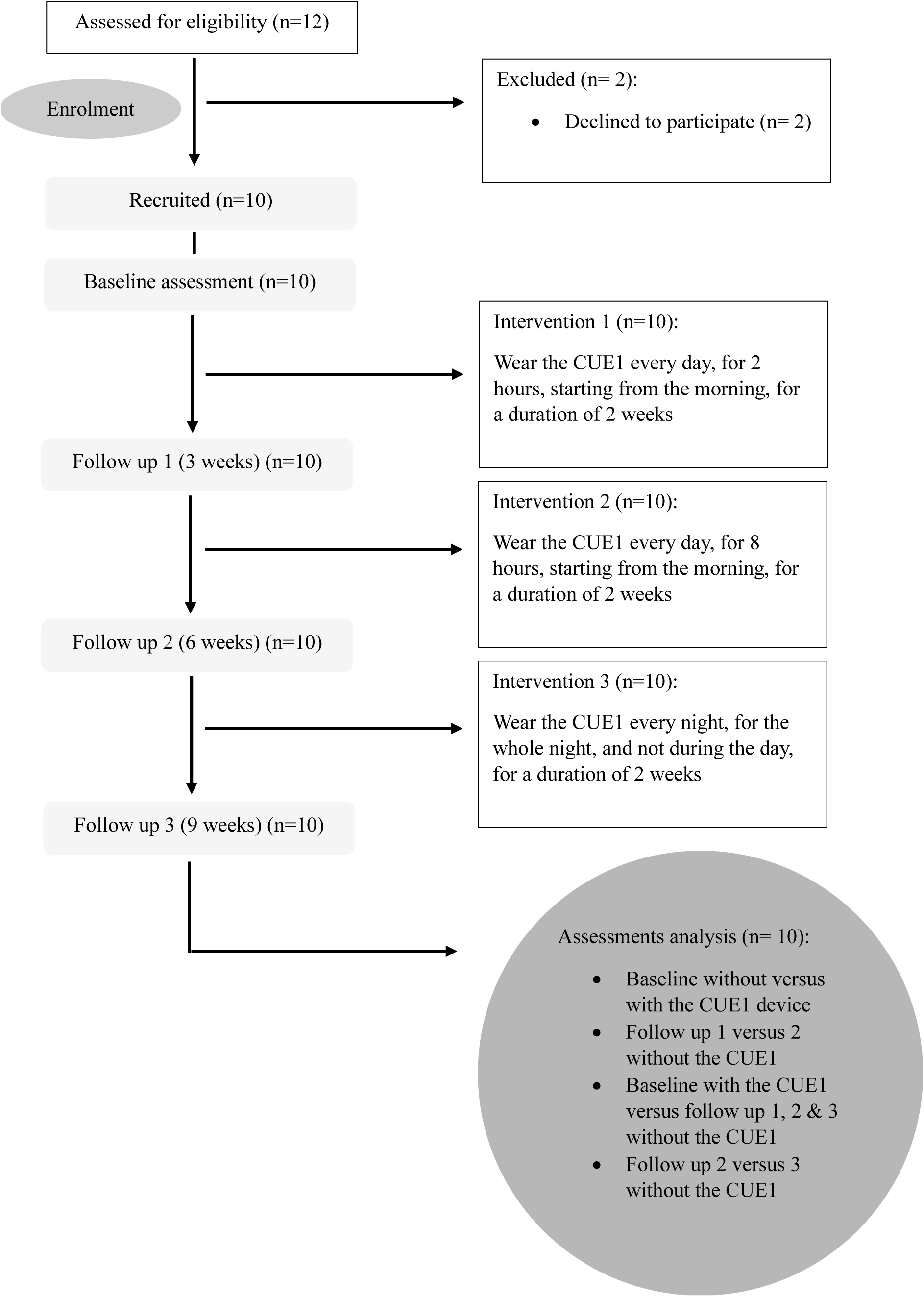
Flow of participants through this study.

Inclusion criteria were clinical diagnosis of PD^17^, adults over 18 years old and willing to participate and provided written consent after reviewing the participant information sheet. Participants were excluded if they had an alternative diagnosis that might affect their movements, balance and ability to carry out the trial independently. These include any neurological disorders other than PD, osteoarticular problems, visual disturbances, audio-vestibular disorders, and cognitive impairments (e.g., a clinical diagnosis of Alzheimer’s disease or Dementia). We also excluded those PwP receiving other non-invasive interventions for PD-related movement symptoms and/or were not on stable PD medication for minimum three months. Moreover, exclusion applied to individuals with implanted metallic or electronic devices, known hypersensitivity to vibrotactile stimulation; and/or skin conditions or open wounds near the device application site (e.g., sternum).

### Measures

Baseline demographic data including age, gender, ethnicity, disease severity (Hoehn & Yahr scale; H&Y)^18^, disease duration, hand most affected by PD, and cognitive status (MoCA) were collected. The MoCA is a screening tool for cognitive impairment, assessing various domains like attention, memory, and executive function^19^. A score above 25/30 suggests normal cognition.

### Outcomes

Feasibility was the primary outcome and was focused on recruitment rate (percentage of eligible participants enrolled), compliance (percentage of days and duration of CUE1 use), and drop-out rates (total number of dropouts).

The secondary outcomes included tolerability and safety. They were assessed through physical examination at baseline, weeks 3, 6, and 9, noting any adverse events related to the adhesive patches. CUE1 usage frequency and duration, comfort and adverse events/reactions concerning both the adhesive patches and the CUE1 device were recorded in participants’ clinical diaries and discussed during appointments. Safety and tolerability were also assessed by using the CUE1 for 2 and then 8 hours during the day and lastly during nighttime.

Effectiveness outcomes were measured at each appointment (baseline, weeks 3, 6, and 9). The objective assessments using the MDS-UPDRS Part-III and subcategories (e.g., Tremor, Bradykinesia, Rigidity, and Gait and Balance)^20^, Functional Gait Assessment (FGA)^21^, and TUG^22^, TUG with dual tasking (DT) (e.g., a cognitive numeracy task; counting backwards from 100 in 7’s)^23^, and Bradykinesia Akinesia Incoordination (BRAIN) tap test^24–26^ were conducted twice: first without the CUE1 and then, after a 20-minute break, with the CUE1 activated on the sternum. All tests were performed in the ON-medication state. Patient-reported outcomes for non-motor features, motor fluctuations, sleep, and QoL were assessed using MDS-UPDRS Part-I, -II, and -IV^20^, Pittsburgh Sleep Quality Index (PSQI)^27^ and Parkinson’s Disease Questionniare-39 (PDQ-39)^28^, respectively (Supplementary material A includes the description of all outcomes).

At the end of the study, patients also completed the participant’s satisfaction form (see supplementary material B) on using the CUE1 device.

### Intervention

This 9-week study involved patients wearing the CUE1 device daily at home while maintaining their usual activities. The CUE1 device^13^ which is Conformité Européene (CE)- marked and regulated in the United Kingdom (UK), is a non-invasive device attached to the sternum via an adhesive patch. All patients used the device with the same settings: 80% vibration strength, 800 ms pulse length, and 800 ms rest length (see image of CUE1 in supplementary material C). The protocol lasted for 9-weeks and was comprised of 3 stages: stage 1: patients wore the device once daily for 2 continuous hours in the morning, starting 1 hour after usual PD medication for first 2 weeks; tage 2: patients wore it once daily for 8 continuous hours in the morning, starting 1 hour after usual PD medication for the next 2 weeks; and stage 3: patients wore the CUE1 only at night for the last 2 weeks (average sleep duration for each participant was obtained based on PSQI questionnaire). Overall participants wore the device for 6 weeks with a week not wearing it in between each stage when the assessments were performed.

### Analysis

The effect of CUE1 was analysed in two ways: the immediate effect was assessed by comparing baseline data for all objective outcomes (MDS-UPDRS Part-III, TUG, TUG DT, FGA, BRAIN) taken on the same day, with and without the device. The cumulative effect was evaluated by comparing baseline data before using CUE1 to follow-up assessments at weeks 3, 6, and 9 without the device, along with questionnaire scores from these time points.

### Statistical analysis

Normality was assessed with the Shapiro-Wilk test, histograms, and Q-Q plots. Wilcoxon signed-rank test identified significant differences on the immediate CUE1 effect. Separate linear mixed effects models were conducted for each motor and non-motor outcome to examine the cumulative effect of the CUE1 intervention within group as a between subject’s factor. Pairwise comparisons were used to locate differences where applicable. All effectiveness outcomes were exploratory and this a p value of 0.05 as cut off score was used. We further considered to use a significance level of p≤0.002 to account for multiple comparisons using Bonferroni correction (e.g., 0.05/24=0.002). Categorical data were presented as absolute numbers and percentages. Analysis was conducted using IBM SPSS version 29 (IBM Corp, Armonk, New York, USA) to obtain descriptive statistics.

## RESULTS

We recruited ten PwP (5 females, age mean ±SD: 66.00±9.83 years; disease duration mean ±SD: 3.90±2.47 years) from diverse ethnic backgrounds (Table 1).

**Table 1.** Patients’ demographic and clinical characteristics.

| Demographic Characteristics | Patients (n = 10) |
| --- | --- |
| Age (years), mean ± SD, range | 66.00 ± 9.83 (46-80) |
| Gender |  |
| Male, n (%) | 5 (50%) |
| Female, n (%) | 5 (50%) |
| Other, n (%) | 0 (0%) |
| Ethnicity |  |
| White or White British (n, %) | 5 (50%) |
| Mixed or multiple ethnic group (n, %) | 2 (20%) |
| Asian or Asian British (n, %) | 2 (20%) |
| Black, African, Caribbean or Black British | 1 (10%) |
| Another ethnic group | 0 (0%) |
| Diagnosis, n (%) |  |
| Parkinson's disease | 10 (100%) |
| Symptom duration (years), mean ± SD, range | 3.90 ± 2.47 (2-9) |
| Symptom severity (H & Y), mean ± SD, range | 2.90 ± 0.57 (2-4) |
| Upper limb most affected by Parkinson's as assessed with MDS-UPDRS III |  |
| Right side, n (%) | 6 (60%) |
| Left side, n (%) | 4 (40%) |
| MDS-UPDRS Part I, mean ± SD, range | 18.60± 6.75 (6.00-26.00) |
| MDS-UPDRS Part II, mean ± SD, range | 17.30± 7.29 (9.00-31.00) |
| MDS-UPDRS Part III, mean ± SD, range | 45.40± 12.22 (26.00-65.00) |
| MDS-UPDRS Part IV, mean ± SD, range | 7.50± 3.75 (1.00-12.00) |
| MDS-UPDRS Total, mean ± SD, range | 88.80± 21.33 (56.00-120.00) |
| MoCA, mean ± SD, range | 26.30 ± 2.50 (23.00-30.00) |

SD, Standard deviation; n, number; %, percentage; H & Y, Hoehn & Yahr; MDS-UPDRS, Movement Disorder Society sponsored revision of the Unified Parkinson's Disease Rating Scale; MDS-UPDRS I: collects information on non-motor aspects of experiences of daily living; MDS-UPDRS II: collects information on motor aspects of experiences of daily living; MDS-UPDRS III: clinically examines motor signs; MDS-UPDRS IV: collects information on motor complications; MoCA, Montreal Cognitive Assessment tool.

### Feasibility, safety and tolerability outcomes

Recruitment rate was high (83%), with excellent compliance (100%) to CUE1 intervention and no dropouts (Table 2). Ten of the 12 participants were recruited; two declined to participate due to time constraints. The CUE1 was safe and well tolerated by all PwP. No side effects were reported. Participants provided positive feedback on their experience with the CUE1 and adhesive patches. All participants reported that they wished to continue using the CUE1 post-trial and would recommend it to other PwP with similar symptoms. Feasibility, safety and tolerability was also explored while the device was used to 2 hours the first two weeks, then 8 hours and finally at night only. These modifications were all feasibility and comfortable for patients. Using the CUE1 for 8 hours daily was more effective than 2 hours, while nighttime did not add additional benefits to motor or non-motor symptoms improvement.

**Table 2.** Feasibility, safety, and tolerability of wearing the CUE1 device over 9-weeks. Information was collected from the participant clinical diary.

| **Outcomes** | **Participants**<br>**(n = 10)** |
| --- | --- |
| **Feasibility** |  |
| Recruitment rate (%) | 10/12 (83%) |
| Compliance with intervention rate (%) | 10/10<br>(100%) |
| Dropout rate (%) | 0/10 (0%) |
| **Safety and Tolerability** |  |
| Total days over the duration of the study of using the CUE1 for each participant | 42 |
| Total number of hours over the duration of the study used the CUE1 for each participant | 240 |
| Total number of side effects arising from CUE1 or adhesive patches | 0.0 (0%) |
| Total number of problems with attaching/removing the adhesive patch for each participant | 0.0 (0%) |
| Average comfort and ease in using the CUE1 device (Likert scale 1-5) | 4.1 |
| Average comfort for feeling/listening the vibration from CUE1(Likert scale 1-5) | 3.7 |
| Average comfort and ease in using the adhesive patch on skin (Likert scale 1-5) | 3.9 |
| Total incidents of CUE1 or charger malfunctioning for each participant | 0.0 |
| **Participant Feedback on Satisfaction with the CUE1** |  |
| How helpful did you find the CUE1 device in relation to your symptoms?<br>(Likert scale 1-5) | 3.4 |
| How satisfied were you with using the CUE1 device? (Likert scale 1-5) | 4.0 |
| If you get the option, how likely are you to continue using the CUE1 device?<br>(Likert scale 1-5) | 4.5 |
| How easy was it for you to use the CUE1 device on the recommended body position (e.g., sternum)? (Likert scale 1-5) | 4.2 |
| How easy was it for you to use the adhesive patches provided with the CUE1 device on the recommended body position (e.g., sternum)? (Likert scale 1-5; %) | 4.1 |
| How likely are you to recommend the CUE1 device to other people with Parkinson's disease and/or related disorder who experience similar symptoms to yours? (Likert scale 1-5; %) | 4.3 |
| How helpful did you find the CUE1 device in relation to your symptoms?<br>(Likert scale 1-5; %) | 3.4 |

N, number; %, percentage; Likert scale where 1 represents the lowest/worst rating and 5 represents the highest/best.

### Immediate CUE1 effect on motor features, gait and falls risk

The MDS-UPDRS Part-III scores showed trends for improvement with the intervention (p=0.008) compared to without it. Trends for improvement were also observed in the MDS-UPDRS Part-III-Rigidity (p=0.008), Bradykinesia (p=0.007), and Balance and Gait (p=0.038). The intervention also resulted in lower (e.g., better) TUG (p=0.022) and TUG DT (p=0.037), and higher (e.g., better) FGA (p=0.007) scores, compared to without the intervention (Table 3). Refer to supplementary material D for BRAIN test scores.

**Table 3.**
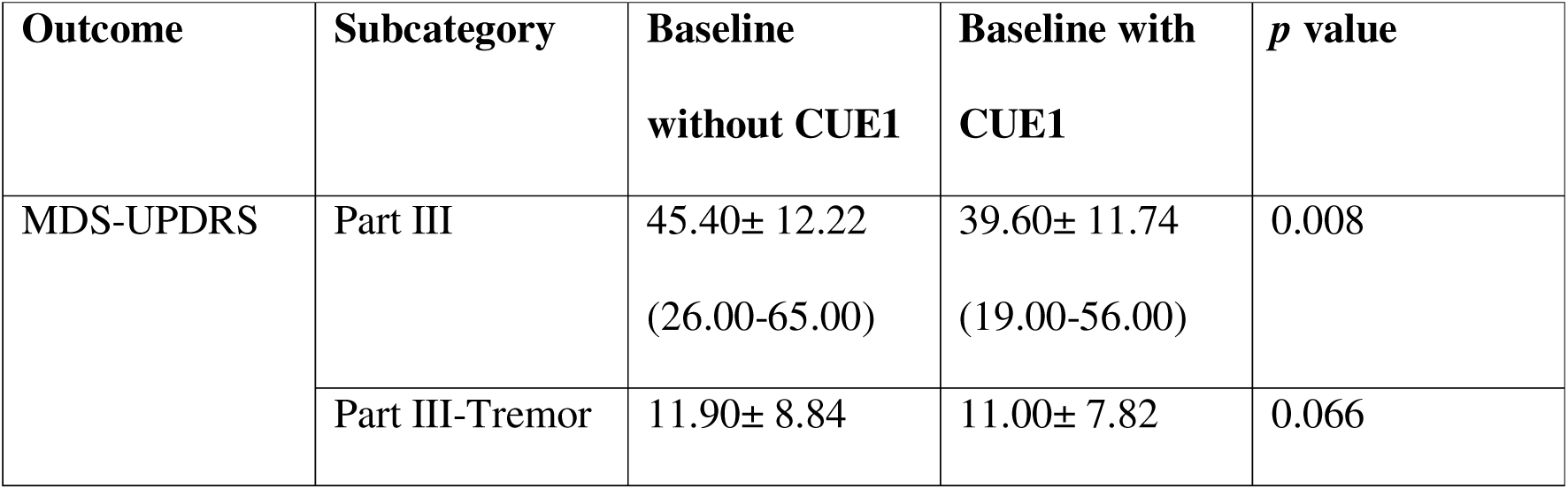
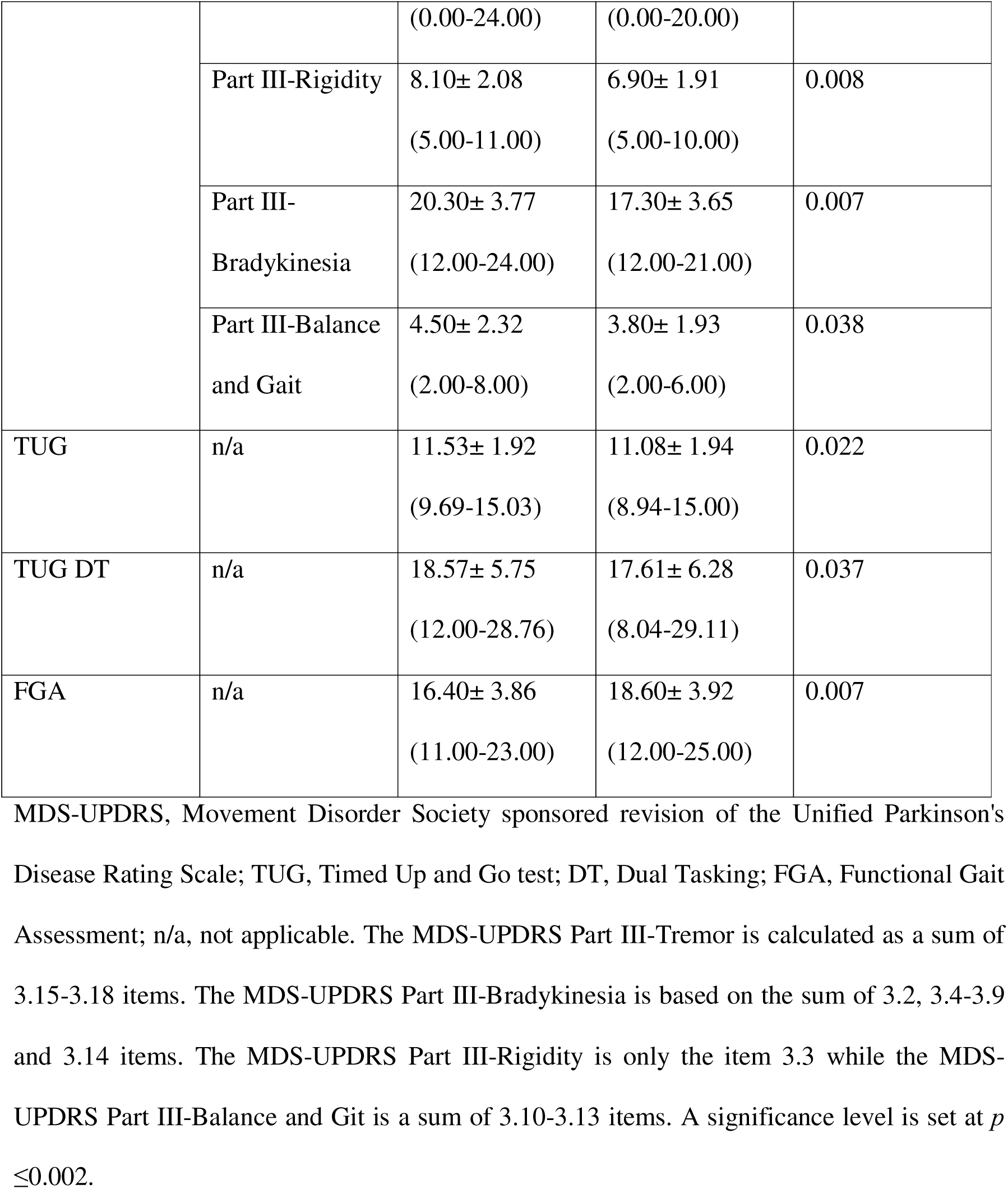
Immediate effect of CUE1 on motor features.

### Cumulative CUE1 effect on motor features, gait and falls risk

There was a positive effect of time (e.g., baseline vs weeks 3, 6 and 9) following the CUE1 intervention on the MDS-UPDRS Part-III [F(3, 21.863)=5.624, p=0.005], MDS-UPDRS Part-III-Rigidity [F(3, 21.873)=4.665, p=0.016], MDS-UPDRS Part-III-Bradykinesia [F(3, 14.282)=4.643, p=0.018], MDS-UPDRS Part-III-Gait and Balance [F(3, 24.279)=4.727, p=0.010], TUG DT [F(3,27.319)=3.429, p=0.031] and BRAIN Incoordination score for the left side [F(3, 15.905)=6.096, p=0.004] (Table 4). The effect of time was significant following the CUE1 intervention on the BRAIN Kinesia score for the left side [F(3, 23.232)=7.717, p<0.001] and FGA [F(3, 25.036)=11.616, p<0.001]. Refer to supplementary material E and F for MDS-UPDRS Part-III, TUG, FGA, and BRAIN test scores, respectively.

**Table 4.**
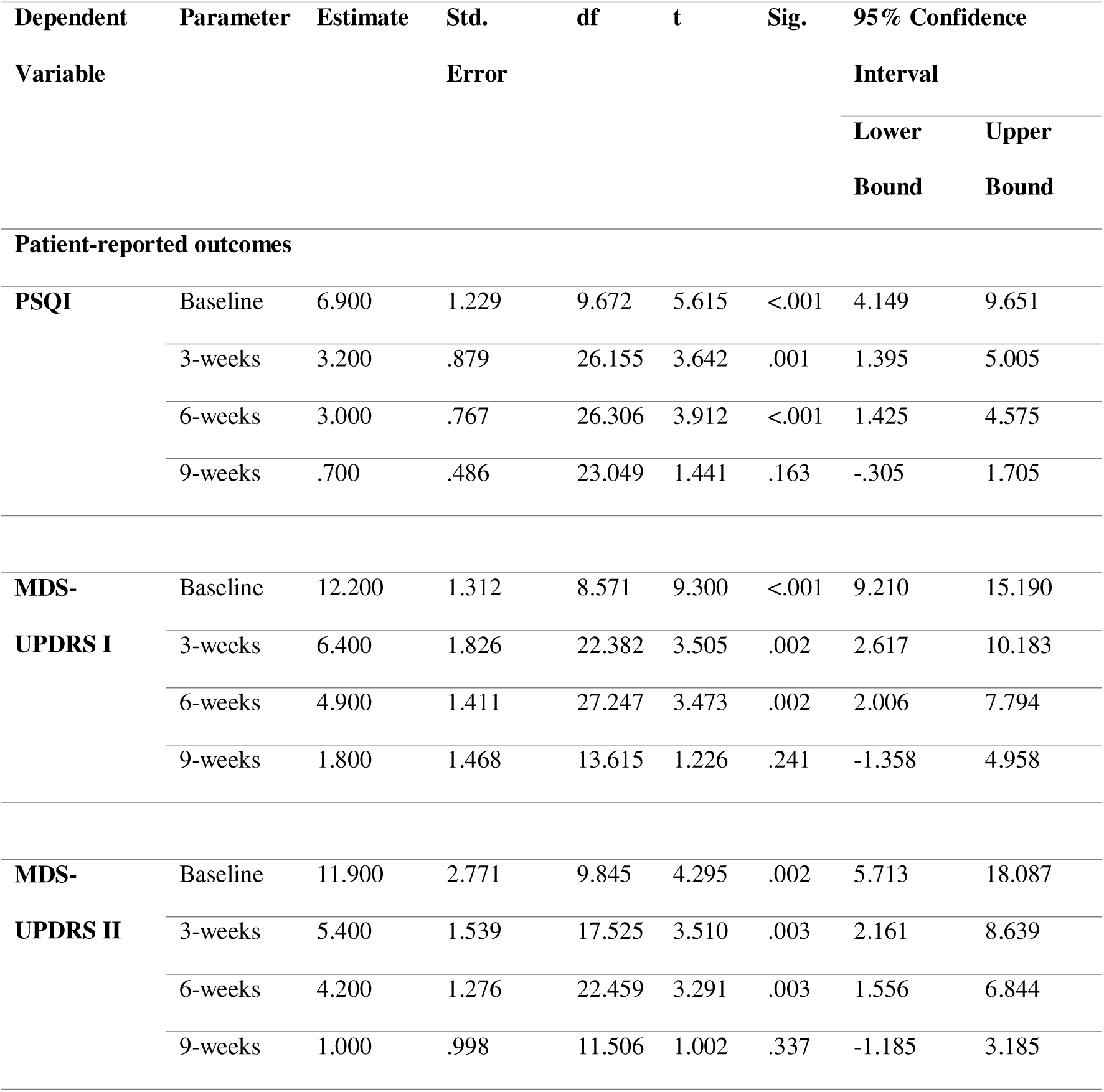
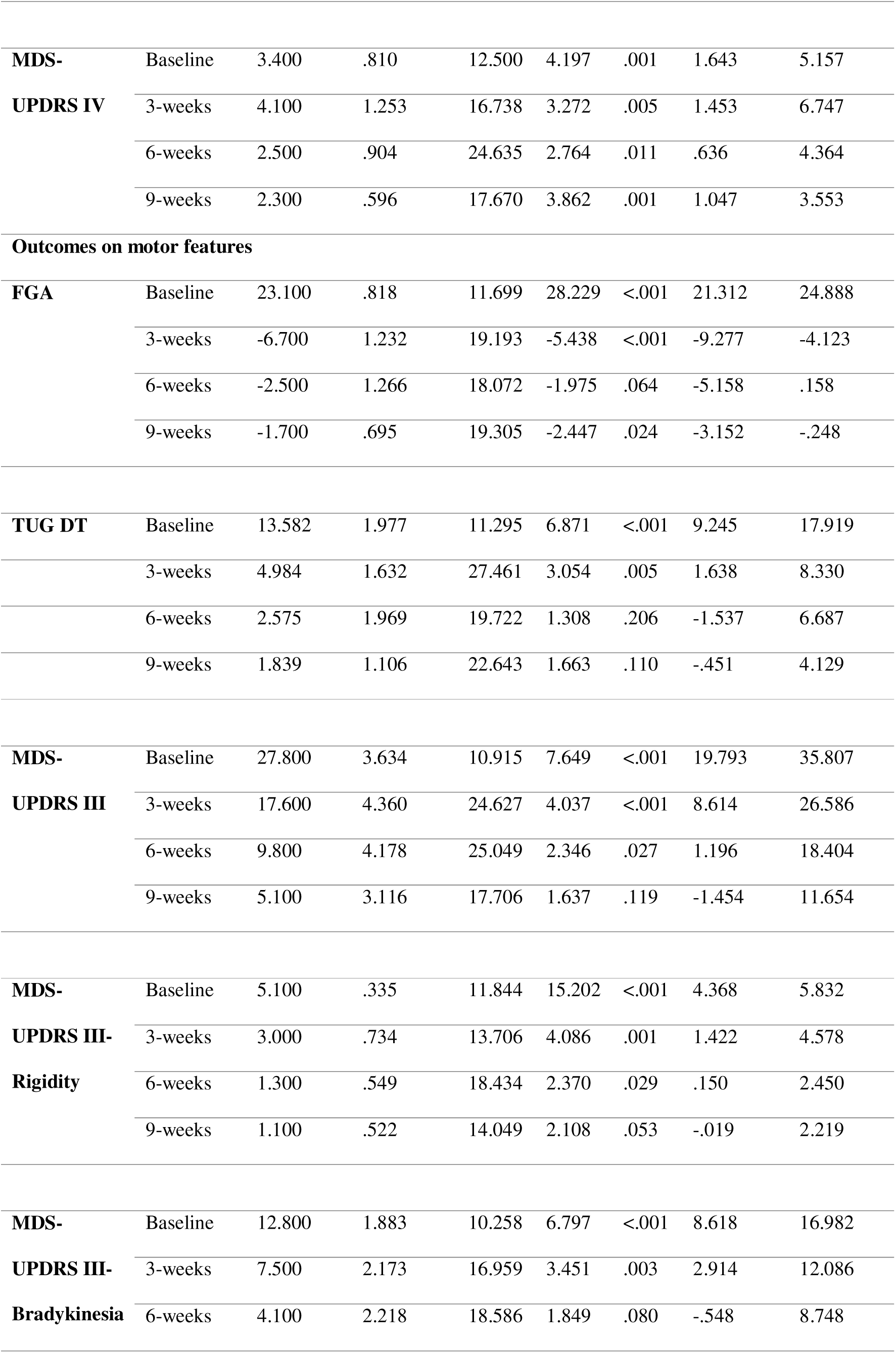
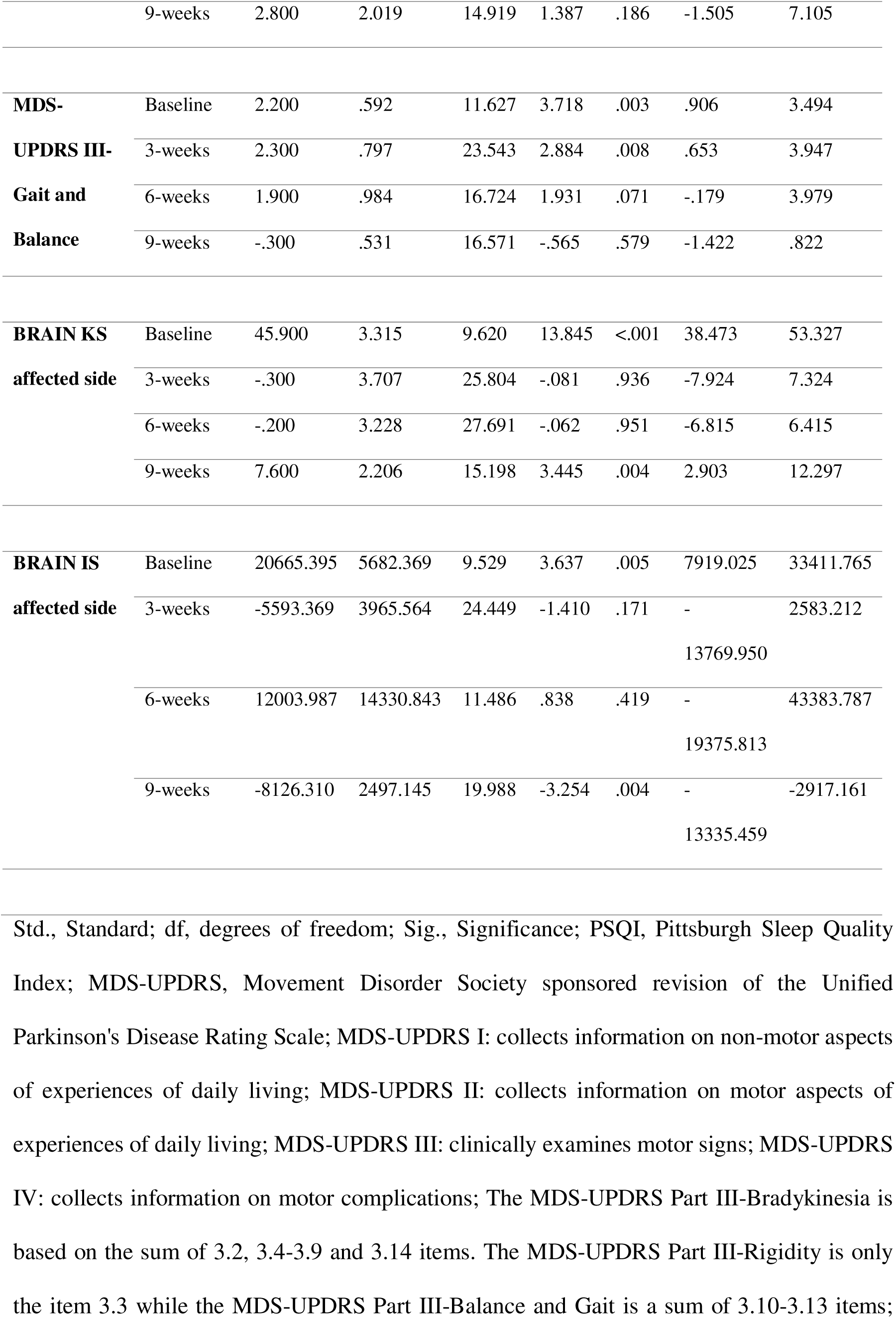
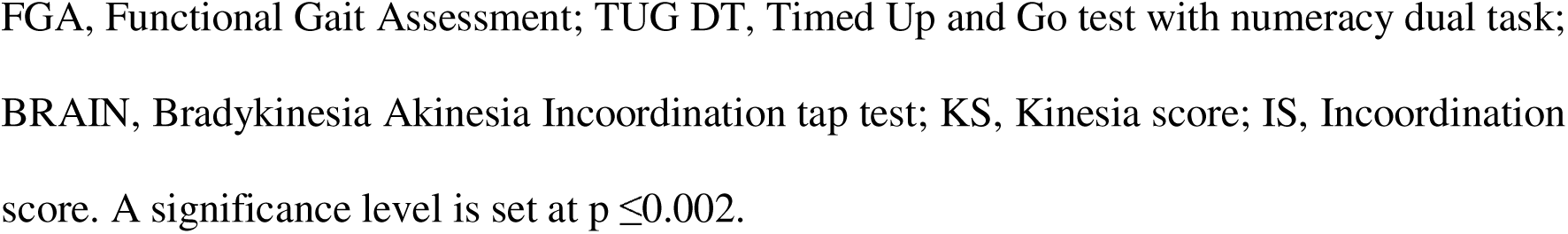
Cumulative effect of CUE1 on non-motor and motor symptoms.

### Cumulative CUE1 effect on patient-reported outcomes

The effect of time was significant following the CUE1 intervention on the PSQI [F(3, 26.131)= 6.275, p=0.002] and MDS-UPDRS Part-II [F(3, 18.226)=7.066, p=0.002] scores (Table 4). There was an improvement following the CUE1 intervention on the MDS-UPDRS Part-I [F(3, 20.358)=4.829, p=0.011] and MDS-UPDRS Part-IV [F(3, 24.482)=6.166, p=0.003] scores. Refer to supplementary material G for patient-reported outcome scores.

## DISCUSSION

This study provides preliminary evidence that cueing with focused vibrotactile stimulation via the CUE1 device is a feasible, safe, and well-tolerated intervention for PwP. The CUE1 intervention was associated with immediate improvements in motor symptoms, along with cumulative benefits for both motor and non-motor symptoms and falls risk reduction. The discussion is divided into sections on a) Feasibility, safety, and tolerability of the CUE1 intervention, b) Effect on motor features, gait and falls risk, c) Effect on patient-reported outcomes, d) Strengths, limitations, and future directions, and e) Conclusions.

### Feasibility, safety and tolerability of the CUE1 intervention

Both cueing and focused vibrotactile stimulation are recognised for their safety and tolerance among PwP^9,10,12–16^. In studies on the Equistasi wearable device, vibrotactile stimulation not only improved motor and non-motor features but also was found to be safe and well tolerated by all patients^12,29^. In this study, the CUE1 device was used over 9-weeks in patients’ homes during their regular routine. Recruitment was successful, probably due to the non-invasive and easy-to-use nature of the intervention. Compliance was excellent, with no dropouts, and no adverse reactions to the CUE1 or its adhesive patches, aligning with user testing results^15^. Wilhelm et al.^15^ noted that the CUE1 reduced motor symptoms and received positive usability feedback, a finding in line with this study. In present study, patients wore the CUE1 for a total of 42 days (e.g., 240 hours), found it comfortable, and reported no malfunctions. Patients rated the overall effectiveness of the CUE1 as 3.4/5 on the Likert scale over 9-weeks and expressed a strong likelihood of continuing use and recommending the CUE1 to PwP with similar symptoms.

### Effect on motor features, gait and falls risk

Some studies indicate that vibrotactile stimulation positively affects performance on MDS-UPDRS III, aiding symptom management in PwP^9,12^. A double-blind randomised controlled trial (RCT) showed significant improvements in MDS-UPDRS Part-III scores after 1 week of using the Equistasi wearable proprioceptive stabiliser, with benefits sustained over 2 months^12^. Similarly, CUE1 intervention led to immediate and cumulative improvements in MDS-UPDRS Part-III scores, including Rigidity and Bradykinesia, over 9 weeks. The immediate reduction was -6.8 points, and the cumulative effect was -11.8, both exceeding the minimal clinically important difference (MCID) of -3.25 points^30^. Another study^16^ reported a 20-point improvement after a 14-week CUE1 intervention in patients with higher H&Y scores. The effect of vibrotactile stimulation on MDS-UPDRS Part-III may vary with disease severity^31^.

The MDS-UPDRS Part-III is a validated semiquantitative scale for assessing motor disability in people with established PD^20^. The scale can distinguish between distal and proximal bradykinesia^32^ and measure the severity of sequence effects and motor arrest. However, the scoring system is accurate enough as there is high interrater variability. The BRAIN tap test, which evaluates amplitude and its decline during hand movements^24–26^, addresses these gaps. While the test requires hand-eye coordination, challenging for visually impaired individuals^26^, our study did not have such patients. Yet, patients may find a challenge to co- ordinate these aspects. We observed cumulative effects of the CUE1 on BRAIN test measures. Although, the speed of movement was slower, the co-ordination was better indicating improved motor control, and precision over 9-weeks. The BRAIN test is effective for remote, long-term upper limb monitoring due to its brevity, ease, and minimal learning curve^26^. Future research should explore the effect of cueing with vibrotactile stimulation on motor control using digital tools like the BRAIN and Digital Finger Tapping^33^, with larger samples to enhance treatment response detection.

Maintaining balance involves coordinating musculoskeletal and neural systems, with input from visual, vestibular, and somatosensory sources^34^. These inputs provide essential information about body position and movement, each offering unique reference points^34^. The central nervous system adjusts its reliance on different sensory inputs based on the task and environment, often increasing dependence on one system while reducing it on others^35^. In PD, the process of challenging reliance on different sensory inputs is impaired, leading to insufficient proprioceptive input and excessive reliance on visual and vestibular cues^36^. Cueing with focused vibrotactile stimulation may help restore this balance during motor planning, improving postural control and gait, as observed in the MDS-UPDRS Part-III-Gait and Balance scores.

This study, consistent with prior user testing, demonstrated that cueing and vibrotactile stimulation improve walking speed and related tasks in both single task^12–14^ and DT^37^ conditions in PwP. The TUG test, commonly used to assess balance and fall risk^38,39^, showed a 7.9 to 256.2 second improvement when using the CUE1 device^13^. Immediate improvements were 0.45 seconds for simple TUG and 0.96 seconds for TUG with DT. Younger and less severely affected patients had different results than Tan et al.^13^. TUG with cognitive DT has a 71% positive predictive value for falls, compared to 42% for simple TUG^40^. Hence, motor tasks with DT should be used to reveal dysfunctions missed during isolated motor tasks^41^, as poor DT performance is a significant predictor of FOG, falls, and reduced independence, impacting QoL in PwP^42^.

About 55% of PwP experience falls annually, with two-thirds having recurrent falls^40^. Cueing and vibrotactile stimulation have been shown to reduce fall risk both acutely and long-term^10,12,29,43^. For example, the Vertiguard device reduced falls by 65.6% and improved body sway^43^. Proprioceptive vibration training decreased fall rates, postural sway area, and anteroposterior axis displacement^29^. An RCT demonstrated a drop in median falls from 4 to 0 during stimulation^12^. The FGA, predicting fall risk with scores ≤18/30^44^, improved from 16.40 to 18.60 with immediate CUE1 intervention and rose to 23.10/30 over 9 weeks, exceeding the MCID of 4 points^45^. This indicates that CUE1 may enhance gait and reduce fall risk, but further research with controls and larger samples is needed. FGA scores also vary between ON- and OFF-medication states, with better predictive validity OFF-medication^46^, suggesting future studies should assess PwP OFF-medication for accurate fall risk prediction.

Subjective improvement in motor symptoms was also supported by improvement seen in MDS-UPDRS Part-III, TUG, FGA and BRAIN test. Improvements in MDS-UPDRS Part-II scores were seen one-week post-intervention and persisted throughout the study. The improvements exceeded the MCID for Part-II (>3.05 points)^47^, with scores increasing by 5.40 points. Additionally, MDS-UPDRS Part-IV improved by 4.10 points, reflecting reduced motor symptom fluctuations. Future research should explore how vibrotactile stimulation impacts medication dosage. An observational study revealed that while levodopa dosage increased in those not receiving vibrotactile stimulation, it remained stable in those who did, suggesting better control of motor symptoms^29^.

### Effect on patient-reported non-motor outcomes

In a double-blind RCT, participants received either focused vibrotactile stimulation plus physical therapy or a placebo device plus physical therapy for 8-weeks^12^. After 9-weeks of CUE1 intervention, notable gains were observed in MDS-UPDRS Part-I, indicating better non-motor daily living experiences. The improvements exceeded the MCID for Part-I (>2.64 points) ^47^, with scores increasing by 6.40 points.

Previous research show that PwP often face sleep issues, such as prolonged bedtime without sleep, trouble staying asleep, and daytime sleepiness^41,47^. Tang et al.^47^ found that high PSQI scores were associated with dyskinesia and motor fluctuations, which aligns with our observation that 90% of PwP had abnormal baseline PSQI scores. Given the link between sleep problems and motor symptoms^41,47^, improvements in motor symptoms may reflect beneficial effect on sleep problems. Despite significant PSQI improvements during the trial, scores remained abnormal post-intervention. The effects of extended or additional sleep-focused treatments on motor symptoms in PwP are not well understood. Further research is needed to explore more sensitive sleep assessments in PwP and to consider integrating sleep therapy with treatments for motor symptoms, such as cueing with vibrotactile stimulation.

### Strengths, limitations and future directions

This study suggests that vibrotactile cueing with CUE1 technology may improve movement symptoms, walking ability, and reduce fall risk in PwP. It could also help with non-motor symptoms and motor complications, but is limited by small sample size, lack of blinding, and short follow-up. Immediate improvements might be influenced by the placebo effect. Future research should use larger samples, longer follow-ups, control groups, and comparisons with sham devices and other treatments. Illness perceptions and cognitive beliefs, which are stronger predictors of quality of life and psychological well-being than clinical factors^48^, should be considered to understand placebo impacts. Given that participants had high MoCA scores, future studies should assess the effect of cueing with vibrotactile stimulation on in PwP with mild cognitive impairment. Daily use of CUE1 for 8 hours was more effective than 2 hours, and nighttime use showed no additional benefit. The one-week washout period questions the sustainability of benefits. Camerota et al.^49^ noted benefits lasting up to 3 weeks, highlighting the need for studies on different usage durations and follow-up periods.

The effects of cueing with vibrotactile cueing on non-motor symptoms in PD are still underexplored^2^. This study assessed the effect of CUE1 on sleep and non-motor symptoms but did not assess separately the MDS-UPDRS Part-I items, thus further research is warrant. Cueing improves cerebellar-cortical activation during walking^50,51^, but its effects on upper limb motor performance require further investigation. Vibrotactile stimulation might reduce beta oscillatory activity in the sensorimotor cortex and enhance motor-sensory connectivity^52,53^. Future studies should validate these mechanisms and address variability in intervention effects. The potential effect of cueing with vibrotactile stimulation should be explored in other conditions such as atypical parkinsonism, multiple sclerosis, Alzheimer’s-related gait disorders, dystonia, essential tremor, and vestibular disorders. A vibrotactile balance belt significantly improved postural control, balance confidence^54^ and QoL^55^ in patients with severe bilateral vestibular hypofunction with many wishing to use it permanently. In a case study, focused vibrotactile stimulation significantly lowered peak neck acceleration during movements and reduced cervical dystonic movements by 60% as demonstrated by electrophysiological and kinematic data^56^.

## CONCLUSIONS

This study offers preliminary evidence that cueing with focused vibrotactile stimulation using the CUE1 device is a feasible, safe, and well-tolerated intervention for PwP. It shows potential for improving motor symptoms, balance, gait, and reducing fall risk, as well as alleviating motor fluctuations, and non-motor symptoms like sleep issues. Future studies with larger sample sizes, control groups and sham device are needed to assess these results.

## Data Availability

The datasets generated during and/or analyzed during the current study are available upon reasonable request from the corresponding authors.

## ACKNOWLEDGEMENTS

We would like to thank all participants for taking part in this study. We appreciate the efforts of all the authors for this article. This study is part of the project funded by Knowledge Transfer Partnership (KTP) United Kingdom 2021 to 2022, round 4, UKRI KTP (Innovate UK) and we would like to thank the organisation for the funding.

## AUTHOR ROLES

VA: 1A, 1B, 1C, 2A, 2B, 2C, 3A, 3B, 3C KR: 1C, 2C, 3C

EC: 1C, 2C, 3C KCD: 1C, 2C, 3C AZ: 1C, 2C, 3C

CQ: 1C, 2C, 3C TB: 1C, 2C, 3C DG: 1C, 2C, 3C

AJN: 1A, 1B, 1C, 2C, 3C CS: 1A, 1B, 1C, 2C, 3C

## DISCLOSURES

### **2)** Financial disclosures for the previous 12 months

Prof Alastair John Noyce reports grants from Parkinson’s UK, Barts Charity, Cure Parkinson’s, National Institute for Health and Care Research, Innovate UK, Virginia Keiley benefaction, Solvemed, the Medical College of Saint Bartholomew’s Hospital Trust, Alchemab, Aligning Science Across Parkinson’s Global Parkinson’s Genetics Program (ASAP-GP2) and the Michael J Fox Foundation. Prof Noyce reports consultancy and personal fees from AstraZeneca, AbbVie, Profile, Bial, Charco Neurotech, Alchemab, Sosei Heptares, Umedeor and Britannia, outside the submitted work. Prof Noyce has share options in Umedeor. Prof Noyce is an Associate Editor for the Journal of Parkinson’s Disease. Dr Cristina Simonet reports grants from Innovate UK and the Michael J Fox Foundation. She also works as a co-investigator with Roche, outside the submitted work.

## ETHICAL COMPLIANCE STATEMENT

This study was approved in the UK by the Dulwich Research Ethics Committee (reference number: 23/PR/1526; clinical.trials.gov ID: NCT06174948). The study adhered to the Declaration of Helsinki. All patients provided written informed consent before data collection.

We confirm that we have read the Journal’s position on issues involved in ethical publication and affirm that this work is consistent with those guidelines.

## Legends for supplemental files

**Supplementary material A.** Description of motor and non-motor outcomes.

**Supplementary material B.** Participant’s satisfaction form.

**Supplementary material C.** The CUE1 device.

**Supplementary material D.** Immediate effect of CUE1 on the Bradykinesia Akinesia Incoordination tap test.

**Supplementary material E.** Cumulative effect of CUE1 on motor outcomes.

**Supplementary material F.** Cumulative effect of CUE1 on Bradykinesia Akinesia Incoordination tap test.

**Supplementary material G.** Cumulative effect of CUE1 on patient-reported outcomes.

