## Supplementary material for "A Pilot Interventional Study on Feasibility and Effectiveness of the CUE1 device in Parkinson’s disease"

**Supplementary material A.** Description of motor and non-motor outcomes

| **Outcome** | **Subcategories** | **Description** | **Scoring** | **Reference** |
|---|---|---|---|---|
| Movement Disorder Society sponsored revision of the Unified Parkinson's Disease Rating Scale (MDS-UPDRS) | Part I | Evaluates non-motor experiences of daily living through 13 questions, divided into Part IA (investigator-assessed) and Part IB (patient self-administered but reviewable by the rater). | Each MDS-UPDRS item is rated on a scale from 0 (normal) to 4 (severe), reflecting the impact of PD symptoms, with higher scores indicating greater severity.<br><br>Part I: Non-Motor Experiences of Daily Living | Goetz et al.[20] |
| | Part II | Addresses motor experiences of daily living with 13 self-administered questions, | | |

| | | | | |
|---|---|---|---|---|
| | | reviewable by the rater. | Score Range: 0-52 | |
| | Part III | Involves a motor examination with 33 scores based on 18 questions, completed by the rater following specific instruction. | Part II: Motor Experiences of Daily Living Score Range: 0-52<br><br>Part III: Motor Examination Score Range: 0-108 | |
| | Part IV | Covers motor complications with 6 questions, integrating patient information and rater observations. | Part IV: Motor Complications Score Range: 0-28 | |
| Functional Gait Assessment (FGA) | Not applicable | A 10-item test evaluating complex gait tasks like walking with head turns, | It scores between 0 and 3 per item, with a maximum of 30 points, | Wrisley et al.[21] |

| | | | | |
|---|---|---|---|---|
| | | walking and turning and climbing stairs. | where higher scores indicate better performance. | |
| Timed Up and Go (TUG) test | | Assesses mobility, balance, walking ability, and fall risks. | The mean time in patients with Parkinson's has been reported between 10.3-14.8 seconds. | Podsiadho et al.[22] |
| Bradykinesia Akinesia Incoordination test (BRAIN) tap test | Kinesia score; KS | Assess upper limb motor function has been validated in patients with Parkinson's and controls. Participants use the index finger of a single hand to alternately | (a) Kinesia score, (KS), the number of key taps in 30 seconds (s)- higher scores indicate better performance. (b) Akinesia time (AT), the mean dwell | Giovannoni et al.[24]; Noyce et al.[25]; Hasan et al.[26] |

| | | | |
|---|---|---|---|
| | Akinesia time; AT | strike the ‘S’ and ‘;’ keys on a standard computer keyboard, as fast and accurately as possible. The test is repeated for the other hand. | time on each key in milliseconds (ms). (c) Incoordination score (IS), the variance of the time interval in milliseconds between keystrokes. For AT, and IS lower scores indicate better performance. |
| | Incoordination score; IS | | |

| | | | | |
|---|---|---|---|---|
| Pittsburgh Sleep Quality Index (PSQI) | Not applicable | Includes seven component scores: subjective sleep quality, sleep latency, sleep duration, habitual sleep efficiency, sleep disturbance, use of sleeping medication, and daytime dysfunction. | Scores range from 0-21 with a higher total score indicating worse sleep quality. In distinguishing good and poor sleepers, a global PSQI score > 5 yields sensitivity of 89.6% and specificity of 86.5%. | Buysse et al.[27] |
| Parkinson's Disease Questionnaire-39 (PDQ-39) | Mobility | A 39-item self-report questionnaire, which assesses how often patients with Parkinson's experience | Dimension score is the sum of scores of each item in the dimension divided by the maximum possible score | Jenkinson et al.[28] |
| | Activities of Daily Living (ADL) | | | |
| | Emotional Wellbeing | | | |
| | Stigma | | | |
| | Social Support | | | |

| | | | |
|---|---|---|---|
| | Cognition | difficulties across the 8 quality of life dimensions and impact of PD on specific dimensions of functioning and well-being. | of all the items in the dimension, multiplied by 100. Lower scores reflect better quality of life. |
| | Communication | | |
| | Bodily Discomfort | | |

**Supplementary material B.** Participant's satisfaction form.

### Participant satisfaction form

**Study title: 'Feasibility, safety, tolerability and effectiveness of the CUE1 device in Parkinson's disease. A 9-week interventional study.'**

Now that you completed your intervention, we would like you to take a bit of time to answer a few questions. Your feedback is important to us as this will help to gain a better understanding of your experience with the research team and using the CUE1 device in this trial. This form has two sections. This form will take approximately 5 minutes to complete.

**Section 1: This section includes questions about the care you received from the research team during the trial. Please select ONLY ONE option for each of the questions below to rate the overall experience.**

| | not at all | slightly | moderately | very | extremely |
|---|---|---|---|---|---|
| How satisfied are you with the support you received from the research team throughout the trial? | ☐ | ☐ | ☐ | ☐ | ☐ |
| How well explained the study process was | ☐ | ☐ | ☐ | ☐ | ☐ |

| | | | | | |
|---|---|---|---|---|---|
| by the research team? | | | | | |
| How professional did you find that the research team was throughout the trial? | ☐ | ☐ | ☐ | ☐ | ☐ |
| How accessible to you was the research team during the trial? | ☐ | ☐ | ☐ | ☐ | ☐ |
| How trustworthy did you feel that the research team was? | ☐ | ☐ | ☐ | ☐ | ☐ |

**Section 2: Questions about the CUE1 device you used during the trial. Please select ONLY ONE option for each of the questions below.**

| | not at all | slightly | moderately | very | extremely |
|---|---|---|---|---|---|
| How helpful did you find the CUE1 device in relation to your symptoms? | ☐ | ☐ | ☐ | ☐ | ☐ |

| | | | | | |
|---|---|---|---|---|---|
| How satisfied were you with using the CUE1 device? | ☐ | ☐ | ☐ | ☐ | ☐ |
| If you get the option, how likely are you to continue using the CUE1 device? | ☐ | ☐ | ☐ | ☐ | ☐ |
| How easy was it for you to use the CUE1 device on the recommended body position (e.g., sternum)? | ☐ | ☐ | ☐ | ☐ | ☐ |
| How easy was it for you to use the adhesive patches provided with the CUE1 device on the recommended body position (e.g., sternum)? | ☐ | ☐ | ☐ | ☐ | ☐ |
| How likely are you to recommend the CUE1 device to other people with | ☐ | ☐ | ☐ | ☐ | ☐ |

| |
|---|
| Parkinson’s disease and/or related disorder who experience similar symptoms to yours? |

**Thank you for completing the participant's satisfaction form.**

**Supplementary material C.** The CUE1 device.

The CUE1 device has been designed to help motor and non-motor Parkinson's symptoms non-invasively. It is a small device, 40 mm in diameter, 11 mm in height, and weighing 17 g. The CUE1 attaches to the sternum with dermatologically tested adhesive patches. The adhesive patches are waterproof. Each adhesive patch can remain up to 14 days on the sternum without the need to change. The CUE1 device is water-resistant but not waterproof. Therefore, it should be removed before showering. The CUE1 uses a silent motor to provide vibrotactile stimulation with a pattern developed through user and clinical testing. It combines focused vibrotactile stimulation and cueing with its specific wave shape and frequency patterns.

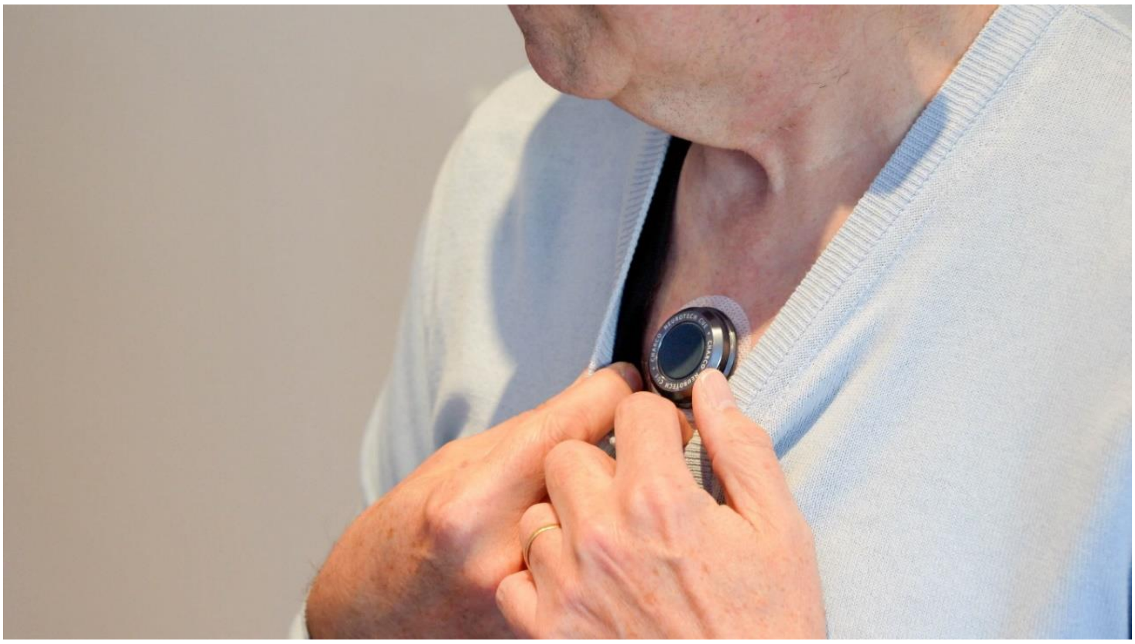

**Supplementary material D.** Immediate effect of CUE1 on the Bradykinesia Akinesia Incoordination tap test.

| Outcome | Sub-category | Side (Left/ Right) | Baseline without CUE1 | Baseline with CUE1 | *p* value |
|---|---|---|---|---|---|
| BRAIN | KS | Left | 43.50± 11.23 (21.00-58.00) | 45.10± 14.39 (14.00-62.00) | 0.610 |
| | | Right | 43.50± 13.32 (12.00-58.00) | 48.20± 13.36 (21.00-67.00) | 0.476 |
| | AT | Left | 155.28± 50.38 (90.09-259.04) | 131.36± 30.75 (86.69-175.24) | 0.285 |
| | | Right | 116.25± 26.85 (87.78-163.90) | 105.71± 15.43 (87.00-136.60) | 0.203 |

| | | | | | |
|---|---|---|---|---|---|
| | IS | Left | 24331.09± 38017.46 (3256.26-131107.40) | 14202.06± 17673.24 (2030.00-62135.21) | 0.074 |
| | | Right | 14965.34± 4977.67 (7947.86-23298.96) | 20423.30± 14149.86 (3714.20-40226.32) | 0.333 |

BRAIN, Bradykinesia Akinesia Incoordination test; KS, Kinesia Score; AT, Akinesia Time; IS, Incoordination Score. A significance level is set at $p \leq 0.002$.

**Supplementary material E.** Cumulative effect of CUE1 on motor outcomes.

| **Outcome** | **Sub-category** | **Baseline assessment** | **3-week assessment** | **6-week assessment** | **9-week assessment** | **P value** |
|---|---|---|---|---|---|---|
| MDS-UPDRS | Part III | 45.40± 12.22 (26.00-65.00) | 37.60± 12.83 (20.00-53.00) | 32.90± 11.57 (18.00-52.00) | 27.80± 12.32 (11.00-48.00) | 0.005 |
| | Part III-Tremor | 11.90± 8.84 (0.00-24.00) | 9.90± 8.24 (0.00-21.00) | 8.90± 7.37 (0.00-19.00) | 7.50± 6.59 (0.00-19.00) | 0.131 |
| | Part III-Rigidity | 8.10± 2.08 (5.00-11.00) | 6.40± 1.65 (3.00-8.00) | 6.20± 2.15 (2.00-9.00) | 5.10± 1.20 (7.00-7.00) | 0.004 |
| | Part III-Bradykinesia | 20.30± 3.77 (12.00-24.00) | 16.90± 4.51 (9.00-22.00) | 15.60± 4.95 (9.00-24.00) | 12.80± 6.27 (4.00-23.00) | 0.018 |
| | Part III-Balance and Gait | 4.50± 2.32 (2.00-8.00) | 4.10± 3.18 (0.00-12.00) | 1.90± 1.97 (0.00-6.00) | 2.20± 2.10 (0.00-7.00) | 0.010 |
| FGA | n/a | 16.40± 3.86 (11.00-23.00) | 20.60± 4.72 (9.00-25.00) | 21.40± 3.31 (16.00-25.00) | 23.10± 2.85 (17.00-27.00) | <0.001 |

| | | | | | | |
|---|---|---|---|---|---|---|
| TUG | n/a | 11.53± 1.92 (9.69-15.03) | 11.84± 4.58 (7.60-23.21) | 10.70± 3.63 (7.71-20.10) | 10.35± 3.50 (7.93-20.05) | 0.255 |
| TUG DT | n/a | 18.57± 5.75 (12.00-28.76) | 16.16± 9.43 (9.07-40.11) | 15.42± 7.62 (8.79-34.17) | 13.58± 7.05 (8.02-33.07) | 0.031 |

MDS UPDRS, Movement Disorder Society sponsored revision of the Unified Parkinson's Disease Rating Scale; FGA, Functional Gait Assessment; TUG, Timed Up and Go test; TUG DT, Timed Up and Go test with Dual Tasking (numeracy task); n/a, not applicable. MDS-UPDRS Part III-Tremor is calculated as sum of items 3.15-3.18; MDS-UPDRS Part III-Rigidity is the item 3.3; MDS-UPDRS Part III-Bradykinesia is calculated as sum of items 3.2, 3.4-3.9, 3.14; MDS-UPDRS Part III-Balance and Gait is calculated as sum of items 3.10-3.13. A significance level is set at $p \leq 0.002$.

**Supplementary material F.** Cumulative effect of CUE1 on Bradykinesia Akinesia Incoordination tap test.

| **Outcome** | **Sub-category** | **Side (Right/Left)** | **Baseline assessment** | **3-week assessment** | **6-week assessment** | **9-week assessment** | ***p* value** |
|---|---|---|---|---|---|---|---|
| BRAIN | KS | Left | 43.50± 11.23 (21.00-58.00) | 39.00± 15.77 (9.00-65.00) | 42.70± 14.29 (10.00-64.00) | 42.10± 12.74 (10.00-59.00) | <0.001 |
| | | Right | 43.50± 13.32 (12.00-58.00) | 47.00± 7.83 (31.00-59.00) | 47.70± 11.85 (18.00-59.00) | 46.80± 11.74 (18.00-61.00) | 0.299 |
| | AT | Left | 155.28± 50.38 (90.09-259.04) | 166.00± 59.46 (97.62-299.69) | 152.09± 41.72 (90.53-219.21) | 146.80± 38.09 (90.53-226.58) | 0.598 |
| | | Right | 116.25± 26.85 (87.78-163.90) | 119.11± 32.80 (95.15-204.90) | 110.73± 17.60 (92.33-146.34) | 114.12± 15.83 (96.76-142.67) | 0.181 |
| | IS | Left | 24331.09± 38017.46 (3256.26- | 23732.95± 26162.16 (6580.39-93906.64) | 9568.72± 7181.73 (2416.74-26995.24) | 14059.91± 9030.96 (4796.96-32746.26) | 0.016 |

| | | | | | | | |
|---|---|---|---|---|---|---|---|
| | | | 131107.40 ) | | | | |
| | | Right | 14965.34± 4977.67 (7947.86- 23298.96) | 12151.55± 9971.25 (970.06- 24775.63) | 18257.85± 14321.12 (1387.47- 38773.05) | 14128.28± 14247.96 (1992.81- 45324.08) | 0.849 |

BRAIN, Bradykinesia Akinesia Incoordination test; KS, Kinesia Score; AT, Akinesia Time; IS, Incoordination Score. A significance level is set at $p \leq 0.002$.

**Supplementary material G.** Cumulative effect of CUE1 on patient-reported outcomes.

| **Outcome** | **Sub-category** | **Baseline assessment** | **3-weeks assessment** | **6-weeks assessment** | **9-weeks assessment** | ***p* value** |
|---|---|---|---|---|---|---|
| PSQI | n/a | 10.10± 4.95 (3.00-21.00) | 9.90± 4.89 (3.00-21.00) | 7.60± 3.78 (2.00-16.00) | 6.90± 3.81 (2.00-14.00) | 0.002 |
| PDQ-39 | Mobility | 138.00± 118.68 (20.00-340.00) | 125.00± 102.53 (20.00-280.00) | 124.00± 96.63 (10.00-290.00) | 124.00± 94.19 (20.00-320.00) | 0.900 |
| | ADL | 126.67± 87.56 (16.67-250.00) | 111.67± 78.98 (16.67-216.67) | 131.67± 90.08 (0.00-300.00) | 108.33± 93.38 (20.00-300.00) | 0.184 |
| | Emotional Wellbeing | 106.67± 72.95 (00.00-216.67) | 100.00± 73.70 (0.00-216.67) | 78.33± 58.29 (0.00-133.33) | 78.33± 52.15 (00.00-133.33) | 0.362 |
| | Stigma | 57.50± 81.69 (0.00-250.00) | 45.00± 65.40 (0.00-200.00) | 45.00± 77.10 (0.00-250.00) | 62.50± 76.60 (0.00-250.00) | 0.532 |
| | Social Support | 90.00± 81.73 (0.00-233.33) | 83.33± 72.44 (0.00-200.00) | 66.67± 44.44 (0.00-133.33) | 73.33± 58.37 (0.00-133.33) | 0.487 |

| | | | | | | |
|---|---|---|---|---|---|---|
| | Cognition | 127.50± 101.00 (0.00-300.00) | 132.50± 97.93 (0.00-300.00) | 145.00± 94.13 (50.00-350.00) | 142.50± 102.10 (25.00-350.00) | 0.683 |
| | Communication | 93.33± 84.33 (0.00-233.33) | 96.67± 82.33 (0.00-233.33) | 70.00± 45.68 (50.00-133.33) | 90.00± 95.65 (0.00-266.67) | 0.382 |
| | Bodily Discomfort | 183.33± 105.70 (0.00-400.00) | 176.67± 106.63 (0.00-400.00) | 133.33± 92.96 (0.00-300.00) | 140.00± 106.32 (0.00-300.00) | 0.050 |
| MDS-UPDRS | Part I | 18.60± 6.75 (6.00-26.00) | 17.10± 5.51 (9.00-26.00) | 14.00± 6.41 (4.00-21.00) | 12.20± 3.68 (7.00-19.00) | 0.011 |
| | Part II | 17.30± 7.29 (9.00-31.00) | 16.10± 8.65 (6.00-32.00) | 12.90± 7.30 (3.00-27.00) | 11.90± 8.67 (2.00-32.00) | 0.002 |
| | Part IV | 7.50± 3.75 (1.00-12.00) | 5.90± 3.25 (2.00-10.00) | 5.70± 2.95 (3.00-10.00) | 3.40± 2.95 (0.00-8.00) | 0.003 |

PSQI, Pittsburgh Sleep Quality Index; n/a, not applicable; PDQ-39, Parkinson's Disease Questionnaire-39; ADL, Activities of Daily Living; MDS-UPDRS, Movement Disorder Society sponsored revision of the Unified Parkinson's Disease Rating Scale; MDS-UPDRS I: collects information on non-motor aspects of experiences of daily living; MDS-UPDRS II: collects information on motor aspects of experiences of daily living; MDS-UPDRS IV: collects information on motor complications; A significance level is set at $p \leq 0.002$.
